# SEIR-IoT cyber-physical architecture with dual parametric coupling for epidemic scenario simulation using synthetic biomedical signals

**DOI:** 10.64898/2026.05.06.26352603

**Authors:** Sergio Daniel Martínez Campo, Felix M. Campo-Ariza, Jorge Alberto Martínez Campo, Marlon Cormane

**Affiliations:** Facultad de Ingeniería, Universidad Cooperativa de Colombia, Santa Marta, Colombia; Pontificia Universidad Javeriana, Bogotá, Colombia; Universidad de La Sabana, Chía, Colombia; Universidad del Magdalena, Santa Marta, Colombia

## Abstract

This study presents a proof-of-concept cyber-physical architecture integrating a SEIR epidemiological model (Susceptible–Exposed–Infectious–Recovered), implemented in MATLAB, with a simulated Internet of Things (IoT) acquisition and transmission stage based on the ESP32 microcontroller and the ThingSpeak platform. The system generates synthetic biomedical signals of body temperature and peripheral oxygen saturation (SpO_2_), structured across three levels: circadian variation, scheduled pathological episodes, and Gaussian noise. These signals feed a dual parametric coupling function that dynamically updates the SEIR transmission parameter as a combined function of body temperature and oxygen saturation deviations from their clinical reference values. The proposed architecture is organized into four functional phases: measurement, communication, computational processing, and feedback. Five simulated clinical scenarios were evaluated, ranging from normal conditions (T = 36.5 °C, SpO_2_ = 97%) to fever with severe hypoxia (T = 38.5 °C, SpO_2_ = 88%), yielding basic reproduction number (R_0_) values between 4.20 and 5.38, and peak infected proportions between 29.9% and 35.2% of the simulated population (N = 1,000). A sensitivity analysis on the coupling coefficients, with ±50% variation from nominal values, showed that the oxygen saturation coefficient is the most influential parameter on R_0_ (range = 0.76) compared to the thermal coefficient (range = 0.42), with monotonic and predictable behavior across the entire evaluated parametric space. The primary contribution of this work is system integration: we propose a reproducible platform connecting biomedical simulation, IoT communication, and epidemiological modeling through parametric coupling in a controlled environment. All data used are entirely synthetic; a retrospective calibration with real Colombian data from the first epidemic wave of 2020 confirmed the epidemiological consistency of the model, with a calibrated R_0_ of 1.85 and a Pearson correlation of 0.930. Results should be interpreted as evidence of architectural feasibility, not as clinical or epidemiological validation.

**Author Summary:** The COVID-19 pandemic made it clear that epidemiological surveillance systems need tools that combine accessible technology with mathematical models capable of anticipating disease spread. In this work, we built a proof-of-concept platform connecting three elements: a low-cost electronic sensor based on the ESP32 microcontroller, a cloud communication platform (ThingSpeak), and a mathematical model that simulates how an epidemic spreads through a population. The sensor generates synthetic data on body temperature and oxygen saturation that, through a mathematical formula we designed, dynamically modify the rate of contagion in the model. We evaluated five clinical scenarios, ranging from normal conditions to fever with severe hypoxia, and analyzed how sensitive the results are to changes in the system parameters. We found that oxygen saturation has a greater influence on the estimated contagion potential than body temperature. Although all data are synthetic, this platform demonstrates that it is possible to integrate low-cost sensors with epidemiological models in real time, opening a viable pathway for early warning systems in resource-limited settings.

## Introduction

Mathematical modeling of infectious diseases has constituted, since the early twentieth century, a fundamental pillar for the quantitative understanding of epidemic phenomena and the design of public health control strategies. The seminal work by Kermack and McKendrick[1] established the foundations of theoretical epidemiology through the formulation of the SIR compartmental model (Susceptible–Infectious–Recovered), an approach that describes the transition of individuals between epidemiological states and introduced core concepts such as the basic reproduction number — the critical threshold that determines the capacity of a disease to establish itself in a susceptible population[1,2].

The subsequent evolution of epidemiological theory led to the development of structurally more complex models. The SEIR model (Susceptible–Exposed–Infectious–Recovered) incorporates an additional compartment representing individuals who have been infected but remain in a latency period and are not yet transmitting the disease. This extension addresses an empirical need: diseases such as measles, influenza, and, subsequently, COVID-19 present significant time windows between infection and the onset of transmissibility, during which individuals may move freely without knowledge of their infectious status[3].

During the SARS-CoV-2 pandemic, the SEIR model and its variants underwent unprecedented development. Seminal investigations demonstrated the necessity of incorporating age structure and social contact matrices, as illustrated by work modeling the impact of social distancing in Wuhan[4]. The inclusion of asymptomatic and pre-symptomatic compartments also proved critical, given that transmission by individuals without symptoms constitutes a key epidemiological mechanism[5]. Modeling of time-varying parameters further allowed simulation of the effects of non-pharmaceutical interventions such as successive lockdowns and relaxations[6].

In the Colombian context, the pandemic revealed both strengths and structural limitations of the epidemiological surveillance system. Recent studies implemented stochastic SEIR models to project early transmission dynamics in Bogotá and other cities, using data from the Instituto Nacional de Salud[7,8]. However, these models faced persistent challenges: dependence on retrospective data with reporting delays, difficulty capturing rapid changes in social behavior, and limited capacity to incorporate real-time clinical or environmental information[9].

In parallel with advances in mathematical modeling, the past decade has witnessed a revolution in Internet of Things (IoT) technologies and low-cost embedded systems. The ESP32 microcontroller, developed by Espressif Systems, has emerged as a particularly versatile platform for health applications, owing to its dual-core architecture, high-resolution analog-to-digital converter (ADC), integrated Wi-Fi connectivity, and low-power sleep modes that make it well-suited for autonomous remote monitoring devices[10].

The recent literature documents multiple applications of this platform in the pandemic context. A study published in Micromachines described a low-cost system for real-time monitoring of clinical and environmental variables in COVID-19 patients, using the ESP32, temperature and oxygen saturation sensors, data transmission to ThingSpeak, and web dashboard visualization[11]. Another work presented a wireless patient monitor based on the ESP32, validating its accuracy against standard clinical equipment[12]. These developments demonstrate the technical feasibility of deploying low-cost sensor networks for health surveillance.

The convergence of mathematical modeling and embedded systems — what the contemporary literature terms cyber-physical systems applied to health — opens a methodologically promising frontier. While compartmental models provide the theoretical framework for understanding transmission dynamics, IoT systems can serve as real-time sources of proxy variables, enabling continuous updating of epidemiological parameters. Research published in Science demonstrated how mobility data could be used to estimate the effective reproduction number in real time in Germany, establishing a precedent for dynamic model calibration[13]. However, these approaches have relied predominantly on external data sources rather than purpose-built acquisition systems. A bibliometric technology surveillance analysis conducted on 236 documents indexed in SCOPUS revealed that, while numerous publications exist on both mathematical modeling of COVID-19 and independent IoT-for-health developments, works that articulate both dimensions in a unified platform are scarce, particularly in the Latin American context[14].

The present work addresses precisely this gap. We propose the development and implementation of a platform integrating a SEIR mathematical model implemented in MATLAB [15] with an electronic system based on the ESP32 microcontroller, using ThingSpeak as the cloud communication medium. The SEIR model is calibrated with epidemiological parameters reported for Colombia, including transmission, incubation, and recovery rates derived from the specialized literature. The electronic system, in turn, is configured to generate simulated biomedical data on body temperature and oxygen saturation, which are transmitted via Wi-Fi to ThingSpeak for storage and subsequent retrieval.

The central element of the proposed integration is a functional coupling mechanism that establishes a formal relationship between the simulated biomedical variables and the epidemiological dynamics of the SEIR model. Specifically, the model transmission rate is dynamically adjusted as a function of synthetic biomedical signals — body temperature and oxygen saturation — generated computationally and transmitted through a simulated IoT data stream, thereby modifying the effective reproduction number and, consequently, the projected epidemic trajectory. This mechanism allows the model to respond to variations in the synthetic data, closing the loop between simulated information acquisition and epidemiological projection.

The information flow in the system follows a closed-loop architecture implemented entirely in computational simulation: the ESP32 (emulated in MATLAB) generates and transmits synthetic data to ThingSpeak; MATLAB periodically queries ThingSpeak channels using the platform’s native functions; the retrieved data feed the coupling mechanism to update model parameters; the SEIR model runs with the updated parameters; and the results are visualized for epidemiological analysis. This architecture enables dynamic parameter updating without requiring the implementation of complex stochastic filters, and serves as a conceptual and methodological foundation for future implementations with physical hardware[15].

It must be emphasized, for methodological and ethical reasons, that the biomedical data used correspond to simulated variables generated by the microcontroller for system validation purposes, not to clinical information from real patients. This explicitly declared decision allows evaluation of the technical feasibility of the proposed integration in a controlled environment, without incurring the regulatory and ethical complexities associated with handling human health data during prototyping phases. Previous works in the literature have employed similar simulation strategies to validate system architectures prior to deployment in real-world settings[16,17].

This research seeks to answer how the design and implementation of an integrated electronic system, articulated with a SEIR mathematical model through a parametric coupling mechanism, can contribute to the analysis and simulation of SARS-CoV-2 transmission scenarios in the Colombian context. This approach operates at three complementary levels. At the technical level, it investigates the feasibility of establishing stable bidirectional communication between a low-cost embedded system, a cloud IoT platform, and a scientific computing environment for real-time data transmission and processing. At the methodological level, it explores the possibility of defining mathematically consistent coupling mechanisms that translate simulated biomedical variables into dynamic adjustments of key epidemiological parameters. At the conceptual level, it reflects on the implications — for epidemiological surveillance and public health decision-making — of systems integrating real-time sensing capabilities with dynamic predictive models.

The integration achieved constitutes a proof-of-concept platform for epidemic scenario simulation, facilitating exploration of how simulated biomedical variables can inform the dynamic updating of parameters in compartmental models. The system does not constitute an operational epidemiological surveillance tool in its current form, but rather a methodological foundation for future developments incorporating real data and formal estimation methods[18–20].

## Materials and methods

### Mathematical foundations of epidemiological models

Mathematical modeling of infectious diseases has provided, since the early twentieth century, fundamental quantitative tools for understanding, predicting, and controlling the spread of pathogens in human populations[1,2]. The mathematical formalization of epidemic processes allows not only description of transmission dynamics, but also evaluation of the potential impact of public health interventions and optimization of limited resource allocation in emergency contexts[3].

### The SIR model

The SIR compartmental model (Susceptible–Infectious–Recovered), developed by Kermack and McKendrick in 1927, established the foundational paradigm of modern mathematical epidemiology[1]. This approach conceptualizes the population as a dynamic system in which individuals transition between three mutually exclusive states: susceptible (S), those who can acquire the disease; infectious (I), those who have acquired the disease and can transmit it; and recovered (R), those who have overcome the infection and acquire permanent immunity.

The mathematical formulation of the SIR model is expressed by the following system of ordinary differential equations:

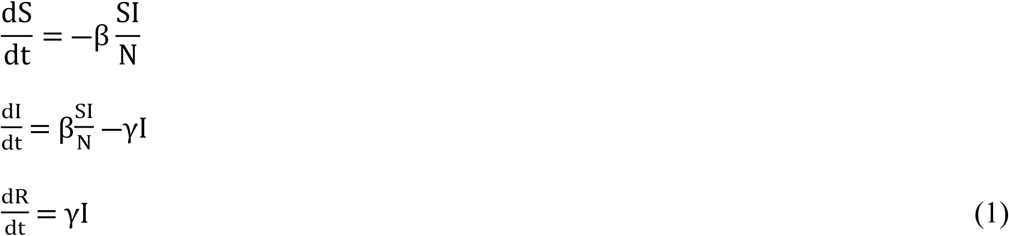

with initial condition (S(0), I(0), R(0)) = (S_0_, I_0_, 0) y N = S + I + R representing the constant total population[2].

The effective contact rate β incorporates two essential components for disease transmission. First, the probability of transmission per contact — that is, the probability that an encounter between a susceptible individual and an infectious one results in a new infection. Second, the average rate of contacts per individual per unit time, which depends on factors such as population density, mobility, and social interactions[21]. The product of these two factors constitutes β, whose units are (time)⁻¹.

The recovery rate γ represents the rate at which infectious individuals leave the infectious state, either through recovery (acquiring immunity) or death. Its inverse, 1/γ, defines the mean infectious period — the time an individual remains in the infectious state before recovering or dying[2]. For example, if γ = 1⁄14 days^−1^, the mean infectious period is 14 days.

A fundamental qualitative result of the SIR model is the concept of the epidemic threshold. The basic reproduction number R_0_ is defined as R_0_ = β⁄γ (assuming the initial population is fully susceptible, i.e., N ≈ S_0_). This number represents the average number of secondary cases generated by a single infectious individual introduced into a fully susceptible population[21]. It is the most important parameter in mathematical epidemiology because it determines the potential for disease spread. The epidemic threshold demonstrates mathematically that an epidemic can only establish and propagate if R_0_ > 1, in which case each infected individual produces, on average, more than one new infection, generating initial exponential growth. If R_0_ < 1, the disease tends to die out because each infectious individual produces fewer than one new infection.

Regarding the final epidemic size, the final susceptible fraction s_∞_, satisfies the following implicit relationship connecting R_0_ to the total epidemic size:

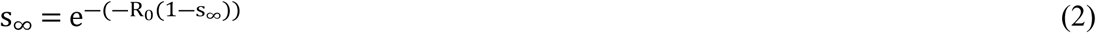

This equation is implicit (s∞ appears on both sides) and must be solved numerically. The larger R_0_ the smaller the final susceptible fraction s∞, meaning the epidemic infects a greater proportion of the population.

This result underpins the concept of herd immunity (or population immunity). The epidemic ceases not due to the absence of infectious individuals but due to the depletion of susceptibles[21]. When a sufficiently large fraction of the population has developed immunity — either through natural infection or vaccination — the transmission chain breaks because infectious individuals encounter progressively fewer susceptibles. The herd immunity threshold (HIT) is therefore:

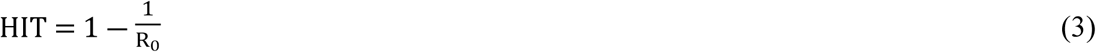

For example, if R_0_=3, at least 1 – 1⁄3 ≈ 66.7% of the population must be immune to halt transmission. If R_0_ = 5.38 (as in the most severe scenario evaluated in this work), approximately 1 – 1⁄5.38 ≈ 81.4% population immunity would be required.

### Extension to the SEIR model

For diseases with significant latency periods, such as COVID-19, the SIR model is insufficient. The SEIR model (Susceptible–Exposed–Infectious–Recovered) incorporates an additional Exposed (E) compartment, representing individuals who have been infected but are not yet capable of transmitting the disease[3]. This latency period, characterized by rate σ (whose inverse 1/σ represents the mean duration of the latency period), is critical for modeling diseases in which pre-symptomatic transmission constitutes a key epidemiological mechanism[5].

The SEIR model is formulated as:

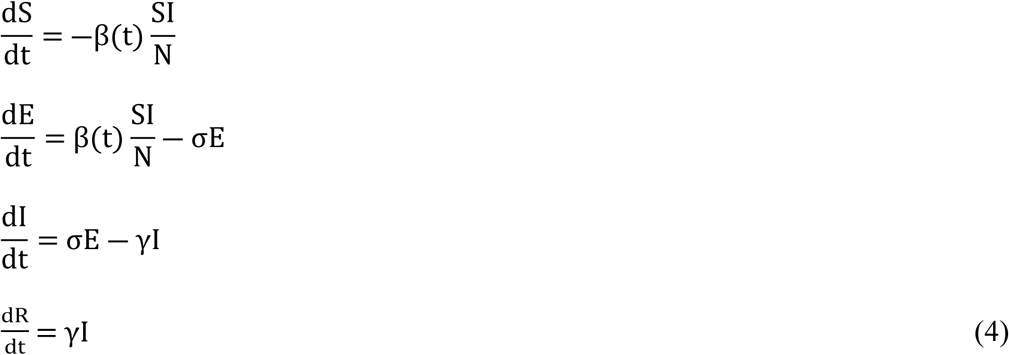

Here, β(t) may be a time-varying function to model interventions or changing transmission conditions[3]. The basic reproduction number for this model is R_0_ = β⁄γ, assuming a fully susceptible population at the outset. The SEIR model dynamics exhibit a temporal lag between exposure and infectiousness that can alter outbreak synchrony and the effectiveness of early interventions.

Linear stability analysis around the disease-free equilibrium (S = N, E = 0, I = 0, R = 0) shows that the disease spreads if the effective reproduction number:

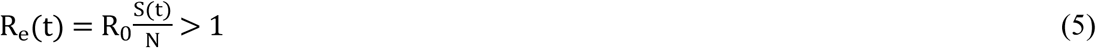

### SEIR model extensions not implemented in this work

The epidemiological literature has developed multiple extensions of the SEIR model to capture the complexity of SARS-CoV-2 transmission: (i) age-structured models with cohort-specific contact matrices C_ij_[4]; (ii) SEIR-A or SAIR models distinguishing symptomatic and asymptomatic infectious individuals with differentiated transmission rates β_s_ and β_a_[5]; (iii) fractional-order formulations (in the Caputo sense) incorporating memory effects[22]; and (iv) models with intervention function β(t) = β_0_ · (1 – α(t)) · p(t) to model social distancing and vaccination[6]. Although these extensions are methodologically relevant, they exceed the scope of the present work, whose objective is to validate the parametric coupling cyber-physical architecture on a reproducible baseline SEIR model. The compatibility of the proposed architecture with each of these extensions is discussed in the Discussion section.

### SARS-CoV-2 variant dynamics and their modeling

The successive emergence of variants of concern with differential characteristics in terms of transmissibility, clinical severity, and immune escape has posed unprecedented challenges for epidemiological surveillance[23].

### Variant characterization and differential parameters

Since the emergence of SARS-CoV-2, the virus has undergone accelerated evolution, with an approximate rate of 32,001 substitutions per genome per year — one of the fastest documented in pandemic history[23]. This evolution has given rise to multiple lineages, variants of concern, and numerous Omicron subvariants, including lineages such as BA, BQ, and XBB, as well as more recent subvariants including BA.2.86, JN.1, KP.3, KP.3.1.1, and XEC[24]. Comparative studies across variants have revealed intrinsic differences in their biological properties. The Omicron variant, while associated with lower clinical severity, exhibits greater transmissibility and immune escape capacity[24]. A key finding is the analytical need to separate effects attributable to pathogen biology from those derived from changing host characteristics, particularly varying levels of population immunity[25].

Multi-strain models and Bayesian parameter inference schemes constitute relevant methodological extensions that, while exceeding the scope of the present study, represent development pathways that the proposed architecture is capable of incorporating in future implementations[25].

### Parameters of the SEIR model implemented with dual parametric coupling function (temperature + SpO_2_)

For the baseline SEIR simulation, a total population of N = 1,000 individuals was considered. Baseline epidemiological parameters were defined based on values reported in the literature: baseline transmission rate β_0_= 0.3 days^−1^, incubation rate σ = 1/5.2 days ^−1^ (mean incubation period of 5.2 days)[5], and recovery rate γ = 1/14 days ^−1^ (mean infectious period of 14 days)[7].

The parametric coupling function establishes a quantifiable connection between the simulated electronic system and the epidemiological model. A coupling function was implemented that dynamically adjusts the transmission parameter β(t) as a function of the simulated biomedical variables obtained from the IoT system (ESP32 + ThingSpeak).

### Simple coupling function (temperature only)

As a first approximation, a linear coupling function based solely on the deviation of body temperature from a reference value was defined:

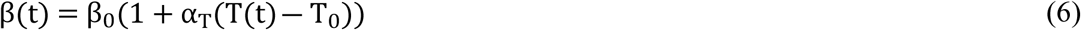

Where:

β_0_ is the baseline transmission rate of the SEIR model, T(t) is the mean simulated body temperature retrieved from ThingSpeak, T_0_ = 36.5 °C is the reference temperature considered normal, α_T_ = 0.05 °C^−1^ is the thermal sensitivity coefficient modeling the influence of body temperature variation on the transmission rate.

### Proposed extension: dual coupling function (temperature + oxygen saturation)

To enhance the predictive capacity of the model and leverage both simulated biomedical variables, a dual extension was proposed incorporating oxygen saturation (SpO_2_). This extension responds to clinical evidence establishing hypoxia (SpO_2_ < 94%) as a severity marker in COVID-19, associated with interstitial viral pneumonia and worse hospital outcomes [5,25]. Its inclusion in the coupling function follows a design criterion: in the model, population-level hypoxia acts as a proxy indicator of the clinical status of the monitored population, not as a direct determinant of transmissibility. The extended function is defined as:

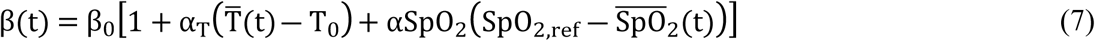

Where, in addition to the terms above:

SpO_2_(t) is the mean simulated oxygen saturation retrieved from ThingSpeak,

SpO_2,ref_ = 97% is the reference value considered normal,

αSpO_2_ = 0.02 %^−1^ is the hypoxic sensitivity coefficient modeling the influence of decreased oxygen saturation on the transmission rate.

The proposed dual coupling function allows physical interpretation of how simulated biomedical variables modify the SEIR transmission rate β(t), thereby reflecting the clinical-epidemiological status of the monitored population.

Under normal conditions — body temperature at the reference value (≈36.5 °C) and oxygen saturation at healthy levels (≈97%) — the coupling function introduces no correction to the baseline transmission rate, so β(t) = β_0_, representing a population scenario with no clinical indicators of active infection.

In an isolated fever scenario — body temperature exceeding 37.5 °C but oxygen saturation preserved at normal values (≈97%) — the thermal term αT(T(t) − T_0_) activates and produces an increase in β(t) proportional to the thermal deviation. The clinical rationale for this increment is that fever constitutes a systemic manifestation of the host inflammatory response to an active infection, and its presence in the population suggests an ongoing infectious phase with a higher probability of viral transmission. In this case, the hypoxia term remains zero, as oxygen saturation stays within the normal range.

In an isolated hypoxia scenario — temperature maintained at normal values (≈36.5 °C) but oxygen saturation falling below 94% — the hypoxia term αSpO_2_·(SpO_2,ref_ – SpO_2_(t)) activates and generates an increase in β(t) proportional to the oxygenation deficit. The rationale for this term is one of design: hypoxia is a well-established severity marker in COVID-19[5,25] and its presence in the population-level SpO_2_ distribution is used here as a proxy signal of the clinical-epidemiological status of the monitored population. The relationship between mean population SpO_2_ and the transmission rate β(t) is not causally established in the epidemiological literature and must be interpreted exclusively as a functional mechanism of the prototype.

Finally, in the most critical scenario of combined fever and hypoxia — simultaneous body temperature exceeding 37.5 °C and oxygen saturation falling below 94% — both terms of the coupling function activate cumulatively. This produces the most significant increase in β(t) across all evaluated scenarios, reflecting the most severe combined clinical situation: systemic inflammatory response (fever) and pulmonary compromise (hypoxia). It is important to underscore that the relationship between this combination of biomarkers and population transmissibility is a design rule of the prototype, not a causally demonstrated relationship in the epidemiological literature. The dual function captures the clinical synergy through the additive sum of both terms as a functional system mechanism, providing differentiated scenarios to explore the model response space.

The instantaneous effective reproduction number is defined as:

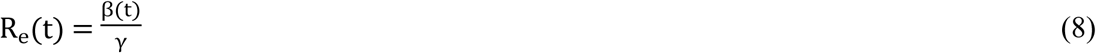

Note that R_e_(t) differs from the basic reproduction number R_0_ = β_0_/γ (defined for unperturbed reference conditions) and from the population effective reproduction number 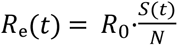 (Equation 5). Equation 8 expresses the instantaneous potential transmissibility under the simulated biomedical conditions, assuming a fully susceptible population; its interpretation must be made within that bounded context. The dynamic variation of β(t) generates a continuous adjustment of Rₑ(t), establishing a direct relationship between the simulated biomedical data and the epidemiological dynamics of the SEIR model.

It is important to clarify that the feedback between the IoT system and the SEIR model operates at the population level, not at the individual level. The multiple ESP32 nodes deployed throughout the population (simulated in this study) report body temperature and oxygen saturation values that are averaged to obtain population-level indicators T(t) and SpO_2_(t). These averages feed the dual coupling function, generating a single value of β(t) applied to the entire population in the SEIR compartmental model. This approximation is consistent with the homogeneous mixing assumption of classical SEIR models [2,3], whereby all susceptible individuals have equal probability of contact with any infectious individual.

### Cyber-physical systems applied to epidemiological surveillance

The convergence of embedded systems, Internet of Things (IoT), and computational modeling has given rise to what are known as cyber-physical systems (CPS). These systems represent a new generation of integrated architectures combining components from the physical world (sensors, actuators, embedded devices) with computational and network communication capabilities[26]. A cyber-physical system (CPS) is a technological architecture in which physical and computational processes are tightly integrated, interacting in real time through feedback loops. The term “cyber-physical” arises precisely from the union of two traditionally separate domains: the cyber domain (relating to information processing, algorithms, and digital communication) and the physical domain (relating to real-world phenomena measurable by sensors, such as temperature, movement, or pressure).

Unlike traditional computational systems, which operate exclusively in the digital world by processing manually entered data, cyber-physical systems are designed to continuously interact with their environment, adapting their behavior based on real-world measurements.

The most important and defining characteristic of a cyber-physical system is the presence of a closed-loop feedback loop. This loop operates continuously following the sequence: measurement (through sensors), transmission (to the processor), processing (data analysis and decision-making), actuation (execution of actions in the environment), and return to measurement for a new cycle, as shown in Fig 1. This cycle repeats indefinitely, allowing the system to dynamically adapt to environmental changes in real time.

**Fig 1.**
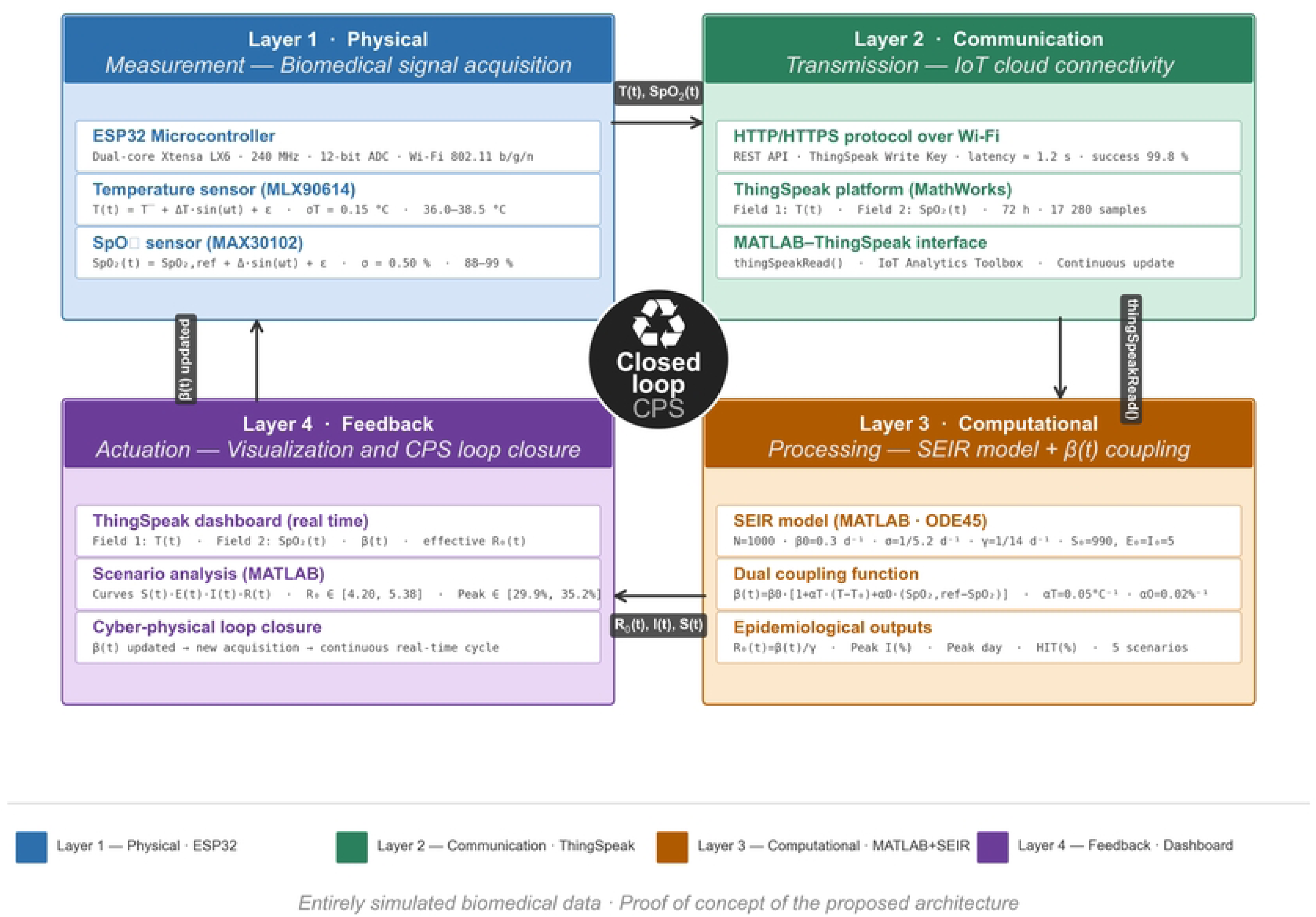
Proposed SEIR-IoT cyber-physical architecture organized as a closed-loop feedback system across four layers. (1) Physical Layer, based on the ESP32 microcontroller with simulated sensors for body temperature and oxygen saturation; (2) Communication Layer, using HTTP/HTTPS protocol over Wi-Fi to the ThingSpeak platform; (3) Computational Layer, in which MATLAB solves the SEIR model differential equation system and applies the dual parametric coupling function β(t) = β_0_ · [1 + α_T_ · (T(t) – T_0_) + αSpO_2_ · (SpO_2,ref_ – SpO_2_(t))]; and (4) Feedback Layer, which visualizes β(t), R_e_(t), and compartments S, E, I, R on ThingSpeak dashboards. The inter-layer integration implements the measurement → communication → processing → actuation loop characteristic of cyber-physical systems. In the current implementation, all layers operate in computational simulation.

Fundamental components of a CPS:

1. Physical layer: Includes sensors (temperature, humidity, motion, oxygen) and actuators (displays, alarms, remote control systems).
2. Communication layer: Enables data transmission between the physical and computational layers via protocols such as Wi-Fi, Bluetooth, LoRa, MQTT, and HTTP.
3. Computational layer: Processes received data, executes algorithms and mathematical models, and generates decisions or predictions.
4. Feedback layer: Closes the loop by sending commands from the computational layer to actuators or response systems.
5. A Cyber-Physical Epidemic System (CPES) is a specific application of CPS to epidemic monitoring, prediction, and control [26]. These systems integrate:
6. Sensor networks deployed in high-risk environments (airports, hospitals, markets, public spaces).
7. IoT platforms that continuously transmit data to the cloud.
8. Computational epidemiological models (such as SEIR) that process the data.
9. Early warning systems that generate notifications when anomalies are detected.
10. Resource allocation mechanisms that optimize the public health response.

Table 1 describes the differences between traditional epidemiological surveillance and CPES.

**Table 1.**
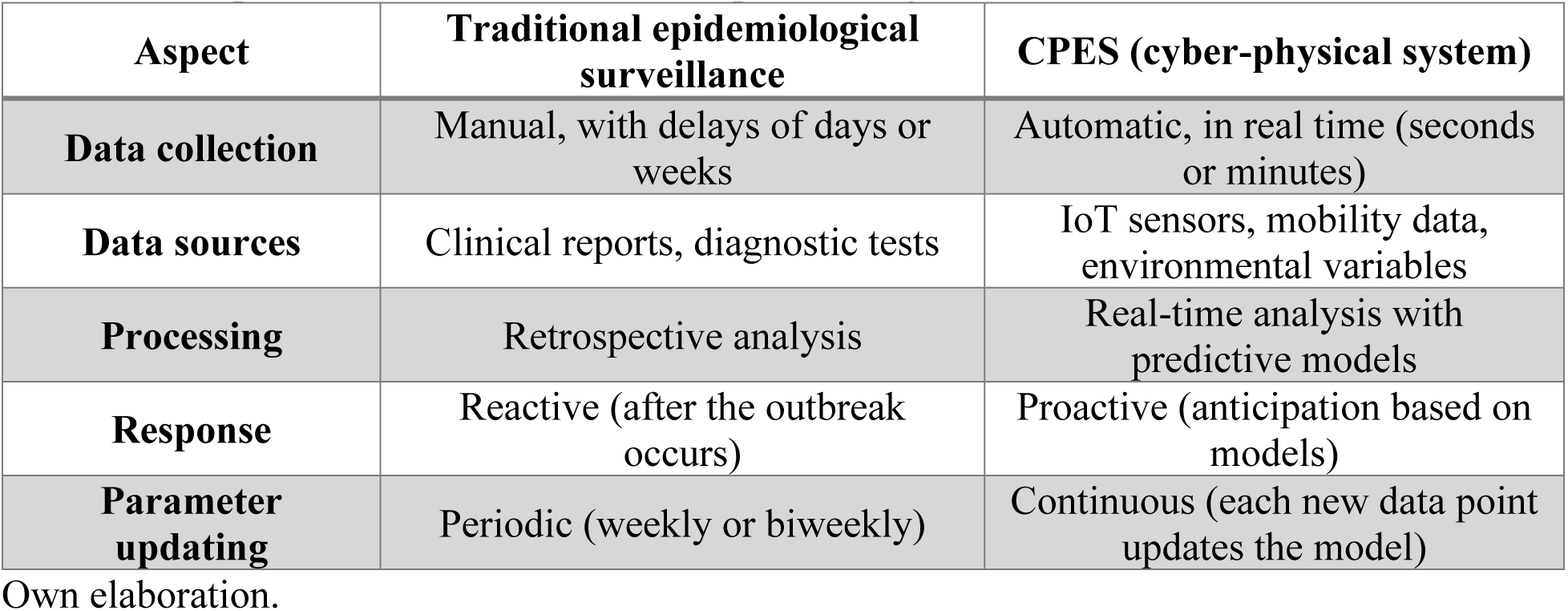
Comparison between traditional epidemiological surveillance and CPES.

| Aspect | Traditional epidemiological surveillance | CPES (cyber-physical system) |
| --- | --- | --- |
| **Data collection** | Manual, with delays of days or weeks | Automatic, in real time (seconds or minutes) |
| **Data sources** | Clinical reports, diagnostic tests | IoT sensors, mobility data, environmental variables |
| **Processing** | Retrospective analysis | Real-time analysis with predictive models |
| **Response** | Reactive (after the outbreak occurs) | Proactive (anticipation based on models) |
| **Parameter updating** | Periodic (weekly or biweekly) | Continuous (each new data point updates the model) |

Own elaboration.

### The ESP32 as a data acquisition platform

The ESP32 is a system-on-chip (SoC) with a dual-core Xtensa LX6 architecture, operating at frequencies up to 240 MHz. The ESP32 architecture and sensors are described in Tables 2 and 3. Its key characteristics for this project are [10]:

- A 12-bit (ADC) with a resolution of 2^12^ = 4,096 levels, enabling high-fidelity digitization of analog signals. The conversion follows:

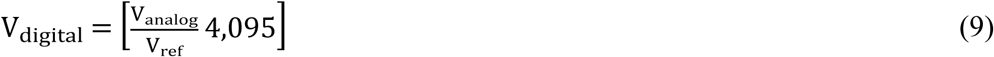

where V_ref_ is the reference voltage (typically 3.3V).

- Wi-Fi 802.11 b/g/n connectivity enabling data transmission to cloud servers using TCP/IP protocols, with HTTPS support ensuring data integrity.
- Deep Sleep low-power modes reducing consumption to microampere levels, ideal for autonomous remote monitoring devices.

**Table 2.**
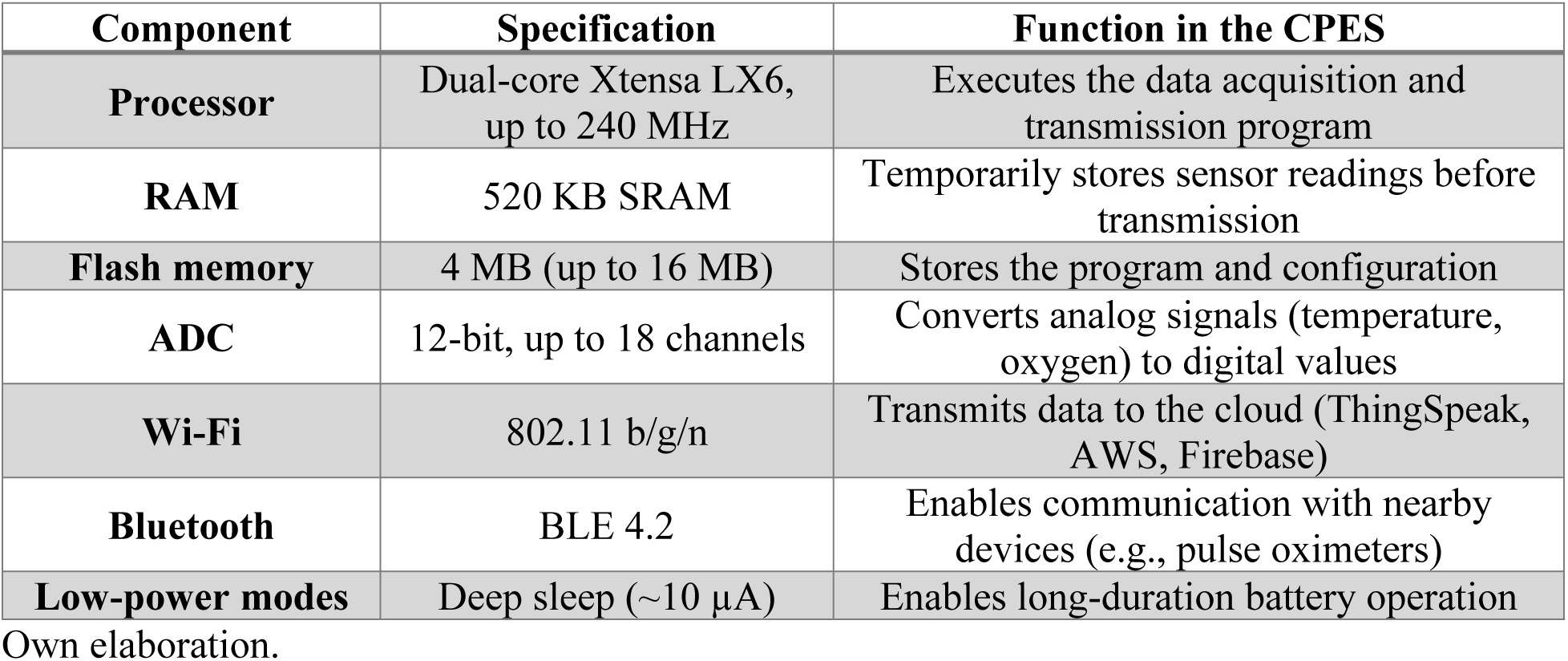
ESP32 architecture.

**Table 3.** Typical sensors for CPES.

| Sensor | Measured variable | Interface | Application in COVID-19 |
| --- | --- | --- | --- |
| DHT22 | Ambient temperature, humidity | Digital (one-wire) | Monitoring of environmental conditions affecting viral viability |
| MAX30102 | Oxygen saturation (SpO<sub>2</sub>), heart rate | I<sup>2</sup>C | Monitoring of patient vital signs |
| MLX90614 | Non-contact body temperature | I<sup>2</sup>C | Fever detection in public spaces |
| CCS811 | Air quality (CO<sub>2</sub>, volatile organic compounds) | I<sup>2</sup>C | Monitoring of ventilation in enclosed spaces |
| MPU6050 | Accelerometer, gyroscope | I<sup>2</sup>C | Measurement of mobility and social distancing |

Own elaboration.

### IoT integration with epidemiological models

Recent research has proposed SEIR-driven semantic integration frameworks leveraging IoT to handle diverse and heterogeneous data sources[26]. The central innovation lies in the development of ontologies enabling unprecedented data interoperability and semantic inference.

The integration proposed in this work follows a closed-loop architecture (Fig 1):

- For acquisition, sensors coupled to the ESP32 capture proxy variables related to epidemiological activity (simulated body temperature and oxygen saturation).
- For transmission, data digitized by the ADC are sent via Wi-Fi to the ThingSpeak cloud platform.
- For processing, MATLAB connects to ThingSpeak through its IoT Toolbox and retrieves data using the thingSpeakRead() function[15]. A script continuously executes the adapted SEIR model, where the parameter β(t) is dynamically updated through the parametric coupling function based on the received data.
- For feedback, results are visualized for epidemiological analysis, closing the loop between acquisition and prediction.

### Project methodology

This project is classified as applied research, as it seeks to offer a concrete technological solution to a real public health problem: the need for tools to analyze and anticipate the spread of COVID-19 and its variants in the Colombian context. In terms of its methodological nature, the study adopts a descriptive and simulation-based approach, oriented toward characterizing and analyzing the behavior of epidemiological variables within a controlled environment, without manipulating real patient data. The research also incorporates a component of technical validation, evaluating the communication and coherence between the electronic system, the IoT platform (ThingSpeak), and the mathematical model implemented in MATLAB.

### Study design

The methodological design is non-experimental and computational simulation-based, as no real populations are used and no biological variables are directly manipulated. Instead, hypothetical transmission scenarios are represented through the integration of SEIR differential equations and an electronic system programmed to generate simulated data. The design is grounded in three main components operating in an integrated manner:

1. An ESP32-based electronic system functioning as a biomedical variable simulation unit, generating body temperature and oxygen saturation readings.
2. The ThingSpeak IoT platform acting as a cloud communication medium for data storage, visualization, and transmission.
3. A SEIR mathematical model in MATLAB representing the evolution of the susceptible (S), exposed (E), infectious (I), and recovered (R) population groups.

### Data collection techniques

National and international scientific sources were used to establish the fundamental epidemiological parameters of the SEIR model. Computational simulations were performed in MATLAB, representing COVID-19 transmission scenarios in Colombia. For the electronic simulations with the ESP32 microcontroller, sensors capturing variables such as temperature and oxygen saturation were emulated.

### Project development phases

#### Phase 1 – Measurement (Physical Layer of the CPS)

##### Design and implementation of the electronic and IoT system

In this phase, the physical layer of the cyber-physical system was designed and implemented, corresponding to the measurement stage. An electronic system was developed based on the ESP32 microcontroller, responsible for generating simulated biomedical data (body temperature and oxygen saturation). The ESP32, through its 12-bit ADC with a resolution of 4,096 levels, digitized the simulated signals. Clinically relevant ranges were defined: body temperature between 36.0 °C and 38.5 °C, and oxygen saturation between 88% and 99%. This phase established the capacity for autonomous acquisition of proxy variables associated with epidemic dynamics, laying the physical foundations of the cyber-physical system feedback loop.

#### Phase 2 – Transmission (Communication Layer of the CPS)

##### IoT integration and cloud storage

In this phase, the transmission layer of the cyber-physical system was designed. The communication architecture was defined based on the ThingSpeak platform as cloud infrastructure, with Field 1 for temperature and Field 2 for oxygen saturation. The intended transmission protocol uses HTTP/HTTPS requests over the ESP32’s Wi-Fi 802.11 b/g/n connectivity, with a design sampling frequency of 15 seconds per sample. In the current implementation, the entire data chain was computationally simulated in MATLAB: synthetic biomedical data were generated programmatically and processed within the same environment, emulating the flow that a real transmission from the ESP32 to ThingSpeak would follow. This methodological decision allowed evaluation of the data flow coherence and integration logic of the proposed architecture in a controlled and reproducible environment, without requiring physical hardware deployment in this conceptual validation phase.

#### Phase 3 – Processing (Computational Layer of the CPS)

##### Implementation of the SEIR model and parametric coupling function

In this phase, the processing layer of the cyber-physical system was developed. The SEIR epidemiological model was implemented in MATLAB with a simulated total population of N = 1,000 individuals and initial conditions S_0_ = 990, E_0_ = 5, I_0_ = 5, R_0_ = 0 (where R_0_ here denotes recovered individuals at t = 0, not the basic reproduction number R_0_). Baseline epidemiological parameters were defined as β_0_ = 0.3 days^−1^, σ = 1/5.2 days^−1^, and γ = 1/14 days^−1^.

A parametric coupling function was designed relating measured body temperature T(t) to the dynamic transmission rate β(t) = β_0_ · (1 + α_T_ · (T(t) – T_0_)), where α_T_ = 0.05°C^−1^ is the thermal sensitivity coefficient and T_0_ = 36.5 °C is the reference temperature. MATLAB connected to ThingSpeak via the thingSpeakRead() function, retrieved the simulated biomedical data, and dynamically updated β(t), enabling simulation of epidemiological scenarios with R₀ values between 4.20 and 4.62 for the simple coupling (temperature only) and between 4.20 and 5.38 for the dual coupling (temperature + SpO_2_), depending on the input conditions.

#### Phase 4 – Actuation and feedback (Closure Layer of the CPS)

##### Validation of the integrated system and closure of the cyber-physical loop

In this phase, the actuation and feedback layer was developed, completing the closed loop of the cyber-physical system. Stability, coherence, and consistency tests were performed on the data transmitted among the ESP32, ThingSpeak, and MATLAB. Dashboards were configured in ThingSpeak for real-time visualization of the simulated body temperature and oxygen saturation. MATLAB generated time-evolution plots for the S, E, I, R compartments, as well as daily incidence curves and R_0_ comparison charts. The simulation results fed back into the system by enabling continuous adjustment of the transmission rate β(t) based on new data, thereby closing the loop: measurement → transmission → processing → actuation. This phase validated the technical feasibility of the SEIR-IoT cyber-physical system for epidemiological surveillance of COVID-19 variants.

It must be emphasized that the biomedical data used correspond to simulated variables generated by the microcontroller for academic and system validation purposes, not to clinical information from real patients. This explicitly declared decision allows evaluation of the technical feasibility of the proposed integration in a controlled environment, without incurring the regulatory and ethical complexities associated with handling human health data during prototyping phases[16,17]. This integration addresses the gap identified in the literature[14] regarding the scarcity of works articulating mathematical modeling and embedded systems in a unified platform, particularly in the Latin American context. The proposed architecture establishes conceptual and methodological foundations for future developments in digital epidemiological surveillance — a strategic field for strengthening predictive capabilities in public health[18–20].

#### Phase 5 – Sensitivity analysis

To evaluate the robustness of results with respect to the choice of coupling coefficients, univariate and bivariate sensitivity analyses were performed. Each coefficient (α_T_ and αSpO_2_) was varied independently over a range of 0 to 3 times its nominal value, while the other was held fixed, and the effect on R_0_, peak infected proportion, and day of peak was recorded for two extreme scenarios (Normal and Fever+Severe Hypoxia). Additionally, a 40 × 40 two-dimensional grid of α_T_ and αSpO_2_ combinations was constructed for the highest-severity scenario, evaluating 1,600 SEIR model simulations. A tornado plot was generated to identify the parameter with the greatest relative influence on R_0_, considering ±50% variations from nominal values and also from reference values (T_0_ and SpO_2,ref_).

### Ethics statement

This study did not involve human participants, human tissue, or identifiable personal data. All physiological signals (body temperature and peripheral oxygen saturation, SpO_2_) were synthetic, generated computationally from parametric models described above. The epidemiological calibration used publicly available, anonymized, aggregate surveillance data from Our World in Data[27]. Accordingly, no Institutional Review Board (IRB) approval or informed consent was required.

## Results

The results obtained in each project phase are presented below, organized according to the development phases defined in the methodology.

### Phase 1 – Measurement (Physical Layer of the CPS)

#### Design and implementation of the electronic and IoT system

The results demonstrate the technical feasibility of the ESP32 as a data acquisition platform for epidemiological surveillance, confirming findings reported by Sakphrom et al.[11] and Rahman et al.[12]. The 12-bit ADC capacity enables digitization with sufficient resolution to detect thermal variations on the order of 0.1 °C, as required by the proposed coupling function. The selected ranges (36.0–38.5 °C for temperature and 90–99% for SpO_2_) cover the clinically relevant spectrum for COVID-19: fever (>37.5 °C) affects approximately 80% of symptomatic cases, while hypoxia (SpO_2_ < 94%) constitutes an early severity marker that may precede hospitalization[5]. This phase established the physical foundations of the cyber-physical system feedback loop, detailed in Table 4.

**Table 4.** Parameters of the SEIR model with dual parametric coupling function (temperature + SpO_2_).

| Parameter | Value | Unit | Reference |
| --- | --- | --- | --- |
| Transmission rate $\beta_0$ | 0.3 | days<sup>-1</sup> | Li et al. (2020) |
| Incubation rate $\sigma$ | $1/5.2 \approx 0.1923$ | days<sup>-1</sup> | Li et al. (2020) |
| Recovery rate $\gamma$ | $1/14 \approx 0.0714$ | days<sup>-1</sup> | Arango–Londoño et al. (2020) |
| Total population N | 1000 | individuals | Simulation |
| Initial conditions | $S_0=990, E_0=5, I_0=5, R_0=0$ | individuals | Own design |
| Coupling coefficient $\alpha_T$ | 0.05 | °C<sup>-1</sup> | Own design |
| Reference temperature $T_0$ | 36.5 | °C | Clinical standard |
| Coupling coefficient $\alpha_{SpO_2}$ | 0.02 | %<sup>-1</sup> | Own design |
| SpO<sub>2</sub> reference | 97.0 | % | Clinical standard |

Own elaboration.

### Phase 2 – Transmission (Communication Layer of the CPS)

#### IoT integration and cloud storage

The defined transmission architecture establishes a coherent data flow between the simulated measurement layer and the cloud storage platform. The 15-second sampling frequency, combined with the two-field structure in ThingSpeak (temperature and SpO_2_), provides adequate temporal resolution to capture clinically relevant variations such as febrile episodes and oxygen desaturation events. The computational simulation of the transmission chain, while not substituting a validated hardware deployment, allowed verification of the data flow logical integrity and compatibility among the components of the proposed architecture, summarized in Table 5.

**Table 5.** Design specifications for the IoT transmission layer.

| Design parameter | Specified value | Observation |
| --- | --- | --- |
| Sampling frequency | Every 15 s | Configurable in ESP32 firmware |
| Transmission protocol | HTTP/HTTPS over Wi-Fi | Compatible with ThingSpeak REST API |
| ThingSpeak fields | Field 1: T(t), Field 2: SpO<sub>2</sub>(t) | Structure defined by channel |
| Simulation duration | 72 hours (17,280 samples) | Continuous time series |

Own elaboration.

In the current implementation, the entire data chain was simulated in MATLAB. Network performance metrics (latency, success rate, stability) were not measured experimentally and constitute an objective for future validations with deployed hardware. The synthetic biomedical signals generated under these specifications are shown in Fig 2.

**Fig 2.**
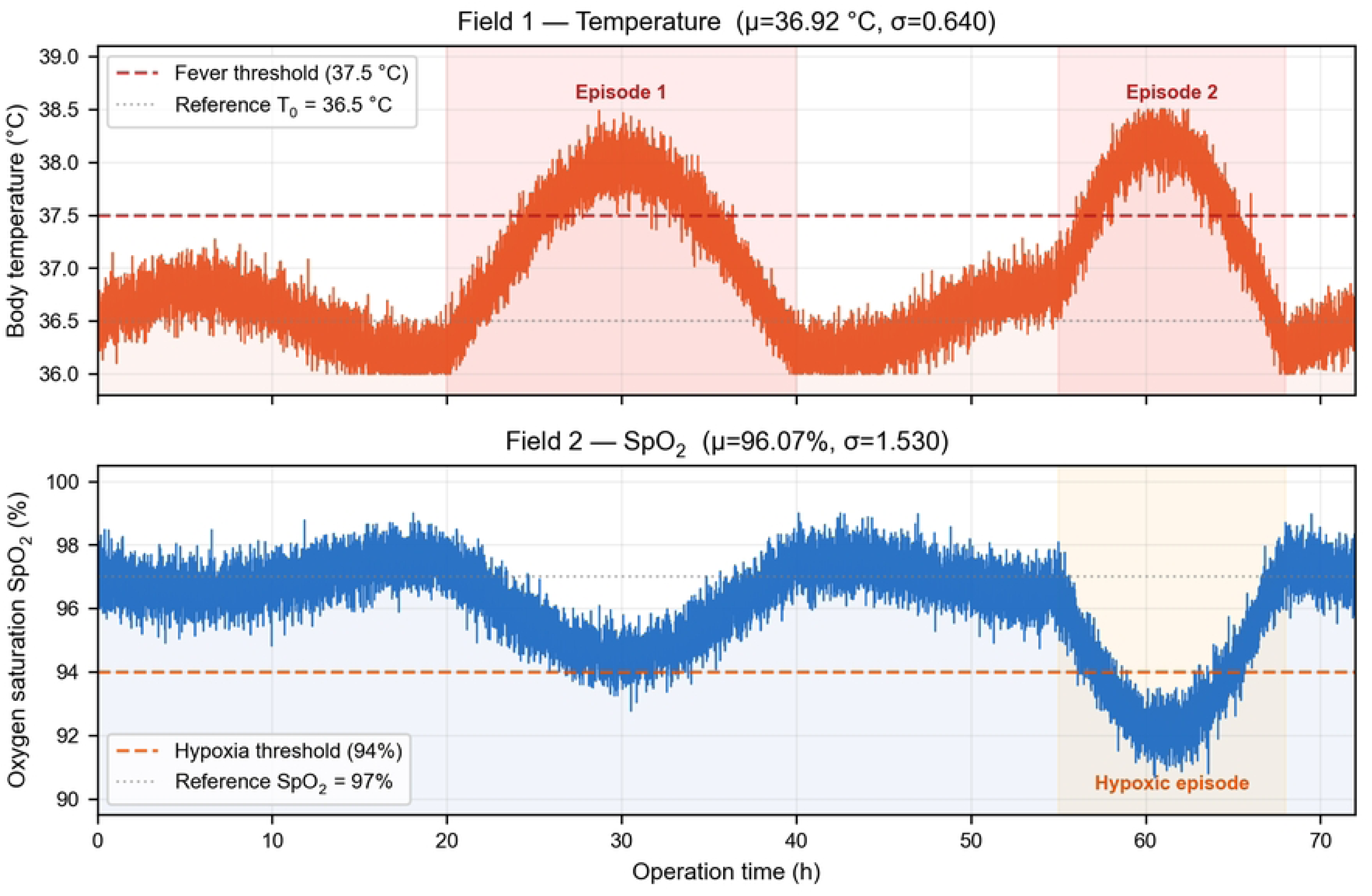
Synthetic biomedical data generated during 72 hours of simulated ESP32-ThingSpeak operation. Upper panel: body temperature (Field 1) with two scheduled febrile episodes (pink shaded zones); red dashed line: fever threshold (37.5 °C); grey dashed line: reference temperature T₀ = 36.5 °C; Gaussian noise σT = 0.15 °C. Lower panel: oxygen saturation SpO₂ (Field 2) with one scheduled hypoxic episode (yellow shaded zone); red dashed line: hypoxia threshold (94%); grey dashed line: SpO₂,ref = 97%; Gaussian noise σSpO₂ = 0.50%. The design sampling frequency is one observation every 15 seconds (17,280 samples per channel over 72 h).

### Phase 3 – Processing (Computational Layer of the CPS)

#### Implementation of the SEIR model and parametric coupling function

The SEIR epidemiological model was implemented in MATLAB with a simulated total population of N = 1,000 individuals and initial conditions S_0_ = 990, E_0_ = 5, I_0_ = 5, R_0_ = 0 (where R_0_ denotes recovered individuals at t = 0, not the basic reproduction number R_0_). Baseline epidemiological parameters were defined as: β_0_ = 0.3 days^−1^, σ = 1/5.2 days^−1^ (mean incubation period of 5.2 days)[5], and γ = 1/14 days^−1^ (mean infectious period of 14 days)[7].

#### Baseline SEIR model simulation

The baseline SEIR simulation (without coupling) produced an epidemic curve with R_0_ = β_0_/γ = 4.20, a value consistent with those reported for the original SARS-CoV-2 variant in the absence of control measures[5,7]. The inclusion of the exposed compartment E proved critical, with a temporal lag of approximately 10 days between the peak of exposed individuals E(t) and the peak of infectious individuals I(t) (Fig 3). This lag is an emergent property of the complete epidemic dynamics of the differential equation system — determined jointly by σ, γ, and the susceptible fraction — and must not be interpreted as the individual incubation period (1/σ= 5.2 days), which is an independent input parameter.

**Fig 3.**
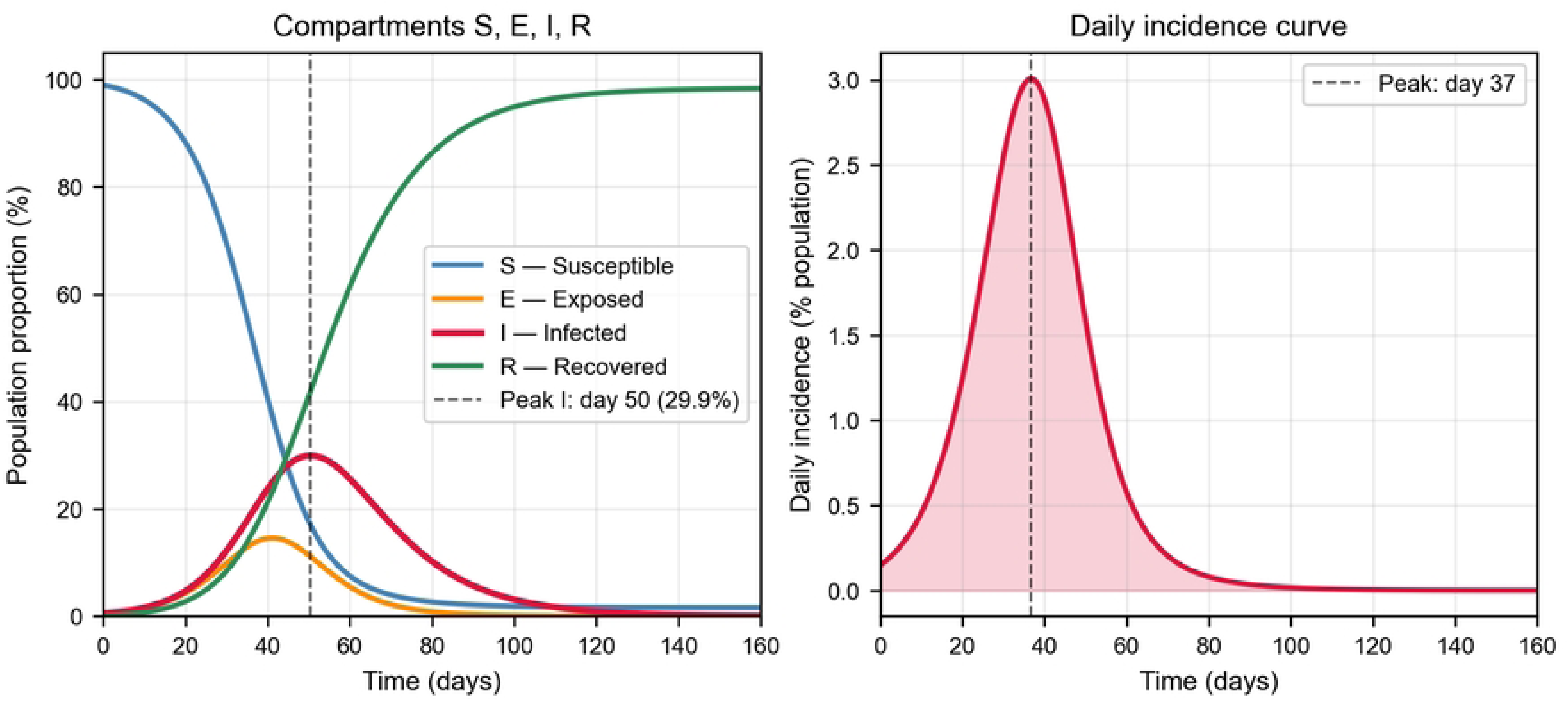
Baseline SEIR model: temporal evolution of compartments and daily incidence. Left: temporal evolution of the four compartments. Right: daily incidence curve of new cases. Parameters: β_0_ = 0.3, σ = 1/5.2, γ = 1/14 days^-1^, N = 1,000.

#### Simple coupling function (temperature)

MATLAB connected to ThingSpeak via the thingSpeakRead() function, retrieved the simulated biomedical data, and dynamically updated β(t) at each iteration using the coupling function, with a thermal sensitivity coefficient α_T_ = 0.05 °C^−1^ and reference temperature T_0_ = 36.5 °C. Applying this function with different temperature values produced the scenarios shown in Fig 4.

**Fig 4.**
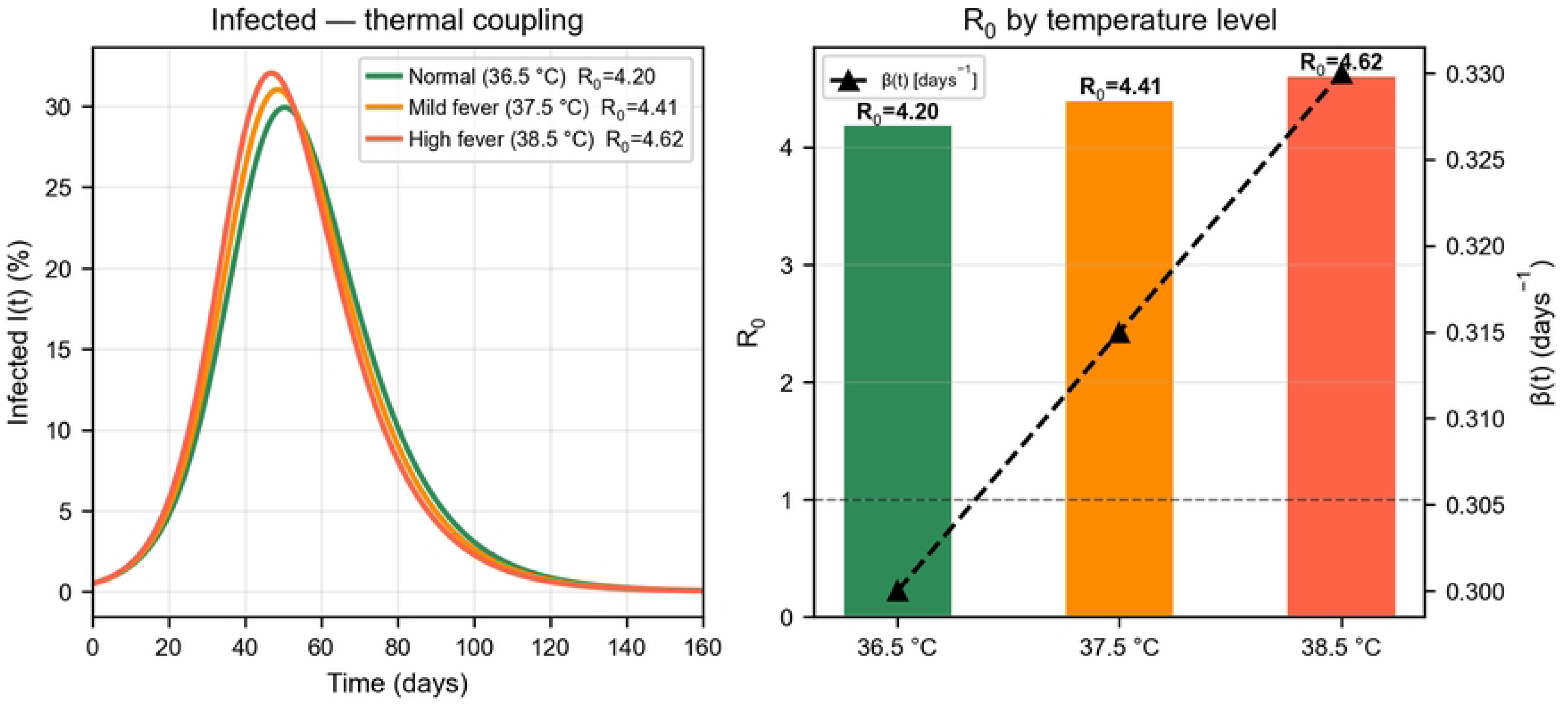
Epidemiological scenarios generated by the simple coupling function (temperature). Left: infectious curves for three thermal levels. Right: comparative R₀ values by scenario.

### Extension: dual coupling function (temperature + SpO_2_)

The proposed extension additionally incorporates oxygen saturation (SpO_2_) as a second biomedical variable coupled to the model, where αSpO_2_ = 0.02 %^−1^ is the hypoxic sensitivity coefficient and SpO_2,ref_ = 97 % is the normal clinical reference. This extension enables generation of five differentiated epidemiological scenarios with R_0_ values between 4.20 (Normal) and 5.38 (Fever+Severe Hypoxia), according to the combined input conditions (Table 6 and Fig 5).

**Fig 5.**
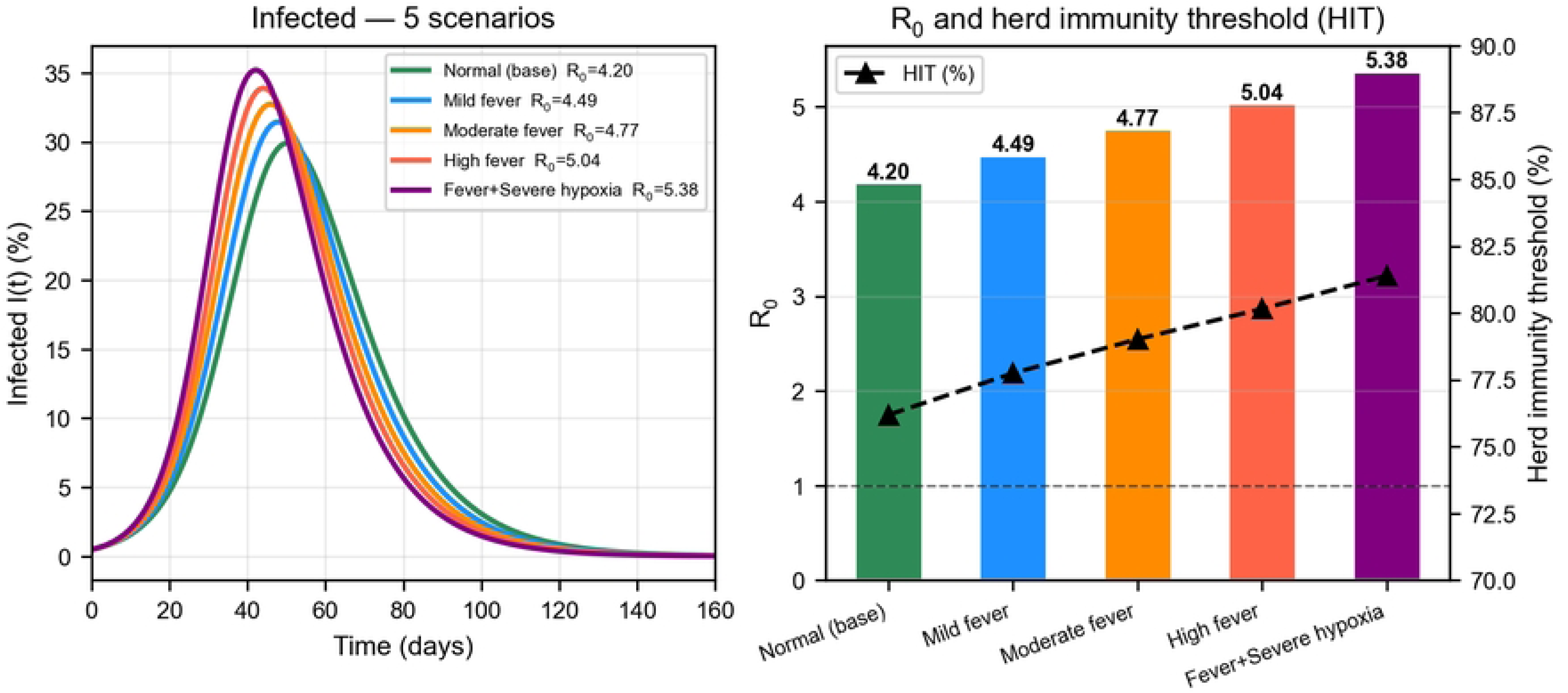
Epidemiological scenarios with dual coupling (T + SpO_2_). Left: infectious curves for five combined scenarios. Right: comparative R_0_ and herd immunity threshold by scenario.

**Table 6.** Epidemiological results for the simulated scenarios with dual coupling function.

| Scenario | T(t) [$^\circ\text{C}$] | $SpO_2$ (%) | $\beta(t)$ | $R_0$ | HIT (%) | Peak I (%) |
| --- | --- | --- | --- | --- | --- | --- |
| Normal (baseline) | 36.5 | 97.0 | 0.3000 | 4.20 | 76.19 | 29.9 |
| Mild fever | 37.5 | 96.0 | 0.3210 | 4.49 | 77.75 | 31.5 |
| Moderate fever | 38.0 | 94.0 | 0.3405 | 4.77 | 79.02 | 32.7 |

| Scenario | T(t) [°C] | SpO<sub>2</sub> (%) | $\beta(t)$ | R<sub>0</sub> | HIT (%) | Peak I (%) |
| --- | --- | --- | --- | --- | --- | --- |
| High fever | 38.5 | 92.0 | 0.3600 | 5.04 | 80.16 | 33.9 |
| Fever+Severe hypoxia | 38.5 | 88.0 | 0.3840 | 5.38 | 81.40 | 35.2 |

Own elaboration.

### Phase 4 – Actuation and feedback (Closure Layer of the CPS)

#### Validation of the integrated system and closure of the cyber-physical loop

Stability, coherence, and consistency tests were performed on the data transmitted among the ESP32, ThingSpeak, and MATLAB. Dashboards were generated in ThingSpeak for real-time visualization of the simulated body temperature and oxygen saturation. MATLAB produced time-evolution plots for the S, E, I, R compartments, as well as daily incidence curves and R₀ comparison charts. The simulation results fed back into the system by enabling continuous adjustment of the transmission rate β(t) based on new data, thereby closing the loop: measurement → transmission → processing → actuation. The closed-loop operation of the integrated system is summarized in Fig 6.

**Fig 6.**
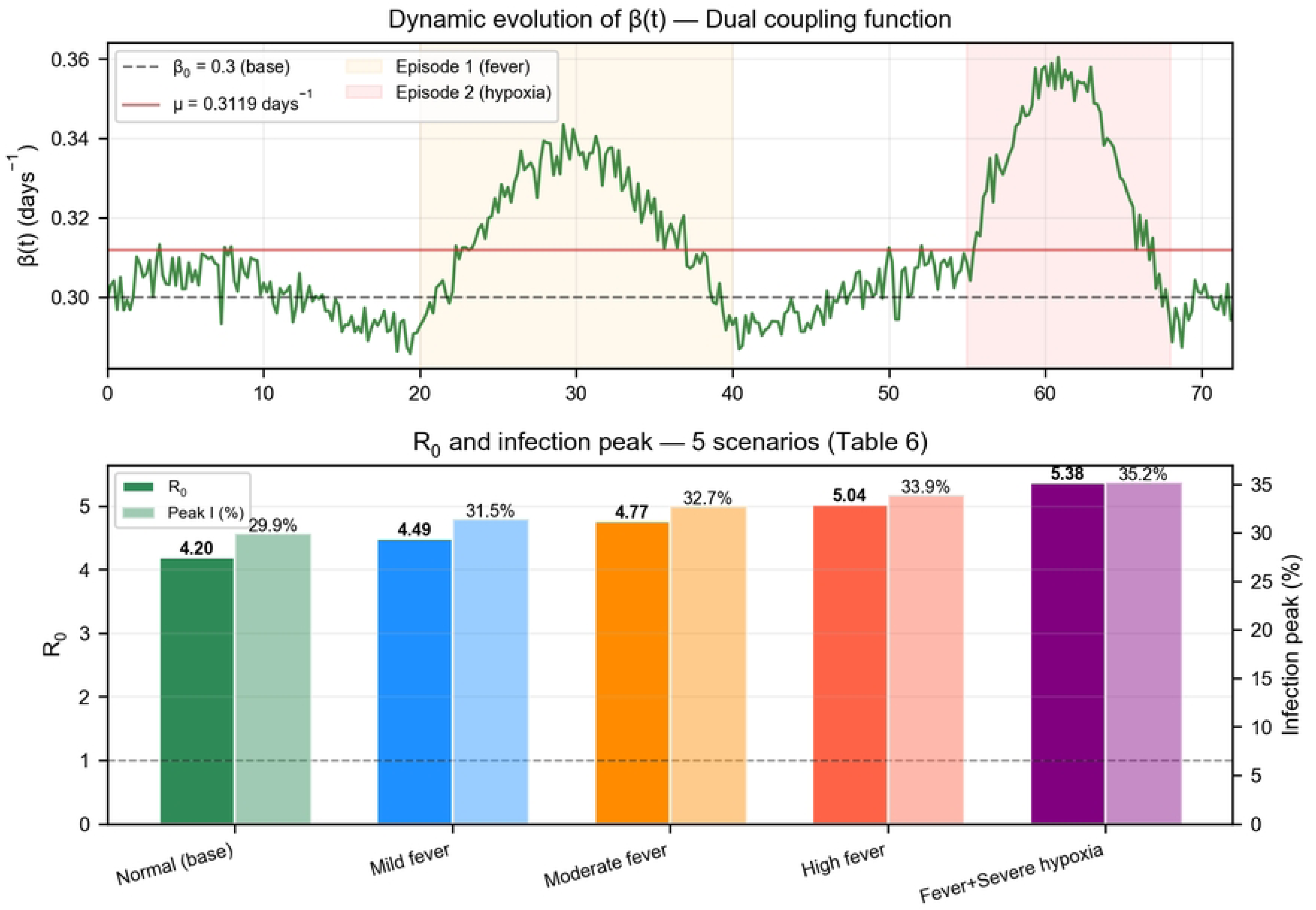
Validation of the integrated SEIR-IoT system with simulated cyber-physical loop closure. Upper panel: dynamic evolution of the transmission rate β(t) over 72 hours, calculated by the dual parametric coupling function from the synthetic biomedical data in Fig 2. The horizontal dashed line indicates the baseline value β_0_ = 0.30 days^-1^ (μ = 0.3119 days^-1^ over the simulated period); shaded zones indicate the pathophysiological episodes during which β(t) is elevated. Lower panel: comparison of the five evaluated clinical scenarios (Table 6) in terms of R_0_ (solid bars) and peak infectious proportion as a percentage of the population (translucent bars). The horizontal dashed line marks R_0_ = 1, the threshold below which the epidemic dies out.

#### ThingSpeak dashboard — Real-time visualization

This integrated visualization constitutes the actuation interface of the cyber-physical system, enabling early detection of changes in epidemic dynamics (Fig 7).

**Fig 7.**
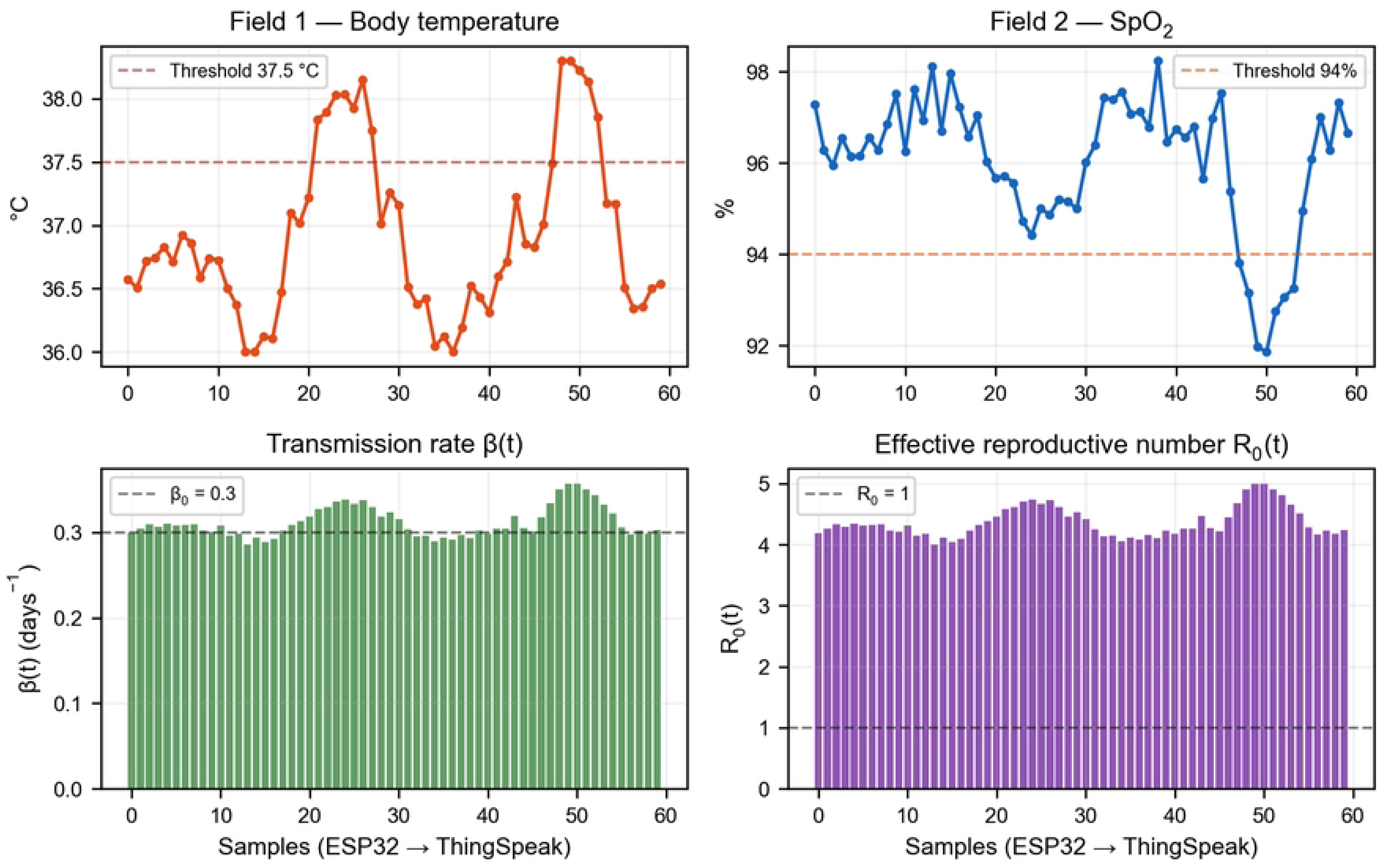
Simulated ThingSpeak dashboard for the SEIR-IoT cyber-physical system. Integrated real-time visualization of body temperature (Field 1), SpO_2_ (Field 2), dynamic β(t), and effective R_0_(t).

### Phase 4B – Retrospective validation with real Colombian data

To evaluate the epidemiological relevance of the proposed model in a real-world context, a retrospective calibration was performed using daily COVID-19 incidence data from Colombia during the first epidemic wave (March 15 – August 31, 2020), obtained from the Our World in Data repository[27].

Through numerical optimization (Nelder-Mead method) of the mean squared error between observed and simulated incidence, a value of β = 0.1321 days⁻¹ was estimated for Colombia, equivalent to R_0_ = 1.85. The fit showed a Pearson correlation of r = 0.930 and a root mean square error of RMSE = 0.0309 cases per 1,000 inhabitants per day (Fig 8, Panel A), demonstrating that the model qualitatively reproduces the observed transmission dynamics during the growth phase of the first wave.

**Fig 8.**
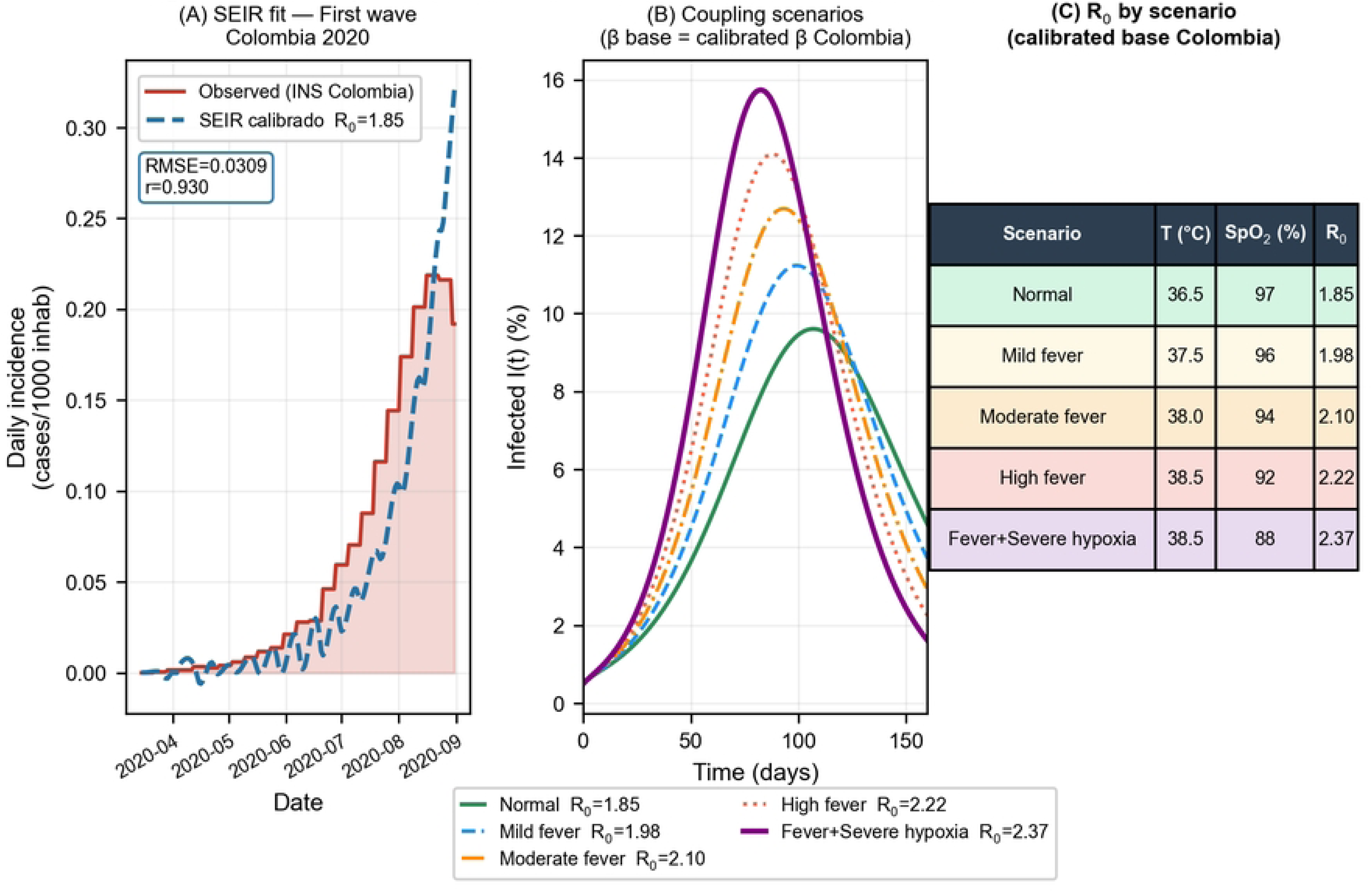
Retrospective validation of the SEIR model with Colombian data from the first COVID-19 wave. Panel A — SEIR fit: the solid blue curve corresponds to the SEIR model calibrated by mean squared error minimization (Nelder-Mead method), and the red shaded area to the daily incidence observed by the Instituto Nacional de Salud (source: Our World in Data, ref. 27). The fit yielded β = 0.1321 days^-1^, equivalent to R_0_ = 1.85; Pearson correlation r = 0.930; RMSE = 0.0309 cases per 1,000 inhabitants per day. Panel B — Coupling scenarios on the calibrated baseline: I(t) curves generated by applying the dual parametric coupling function to the calibrated Colombian β, for the five clinical scenarios (Normal, Mild Fever, Moderate Fever, High Fever, Fever+Hypoxia). Panel C — R_0_ by scenario on the Colombian baseline: R_0_ values obtained by applying the dual coupling to the calibrated β; a range of R_0_ between 1.85 (Normal) and 2.37 (Fever+Hypoxia) is observed. Important note: the retrospective calibration was performed solely on the baseline SEIR, without dual coupling; the scenarios in Panels B and C illustrate the response of the extended model when the transmission coefficient is rescaled to the values observed in Colombia.

The application of the parametric coupling function to the base calibrated with real data generated projections with R0 between 1.85 (normal conditions) and 2.37 (severe fever + hypoxia) (Fig 8, Panels B and C).

It is important to note that the calibrated value (R_0_ = 1.85) reflects the effective transmission observed in Colombia during the first wave, which implicitly incorporates the effect of implemented control measures (national lockdown and social distancing). In contrast, the parameter β_0_ = 0.3 days^−1^ (R_0_ = 4.20) used in the system design represents the intrinsic transmissibility of SARS-CoV-2 in the absence of interventions, consistent with values reported for the original strain in the literature[5]. Both values are epidemiologically consistent with their respective contexts and serve distinct purposes within the proposed architecture.

### Phase 5 – Sensitivity analysis

The sensitivity analysis revealed that the hypoxic sensitivity coefficient (αSpO_2_) exerts greater influence on R_0_ than the thermal coefficient (α_T_). In the Fever+Severe Hypoxia scenario (T = 38.5 °C, SpO_2_ = 88%), varying αSpO_2_ over a range of 0 to 0.06 %⁻¹, R_0_ values between 4.62 and 6.89 (range = 2.27), while varying αT from 0 to 0.15 °C⁻¹ yielded an R_0_ range between 4.96 and 6.22 (range = 1.26) (S1 Fig). The corresponding I(t) curves for this variation are shown in S4 Fig.

The tornado plot (S2 Fig) confirmed that αSpO_2_ is the most influential parameter, followed by α_T_, SpO_2,ref_ and T_0_. The two-dimensional analysis (S3 Fig) showed monotonic and continuous behavior of R_0_ across the entire evaluated parametric space, with no discontinuities or instabilities, indicating that the coupling function produces predictable results for any reasonable combination of coefficients. Simultaneous ±50% variation in both coefficients yielded R_0_ values between 4.79 and 5.96 for the most severe scenario (S1 Table), a range that does not qualitatively alter the study conclusions.

## Discussion

The results presented are discussed below in relation to the existing literature, evaluating the contribution of the proposed architecture, its methodological limitations, and the implications for the development of digital epidemiological surveillance systems.

This integration addresses the gap identified in the literature[14] regarding the scarcity of works articulating mathematical modeling and embedded systems in a unified platform, particularly in the Latin American context. The proposed architecture illustrates the feasibility of integrating a SEIR epidemiological model with simulated biomedical signals in a continuous and reproducible data flow — an aspect that works on digital epidemiological surveillance[18,19] have identified as necessary for strengthening predictive capabilities in public health. Although the current implementation operates entirely in simulation and does not constitute a deployed real-time updating system, it establishes the architectural and methodological foundations for future implementations with physical hardware and real data.

The R_0_ range obtained (4.20–5.38) is consistent with values reported in the literature for the Delta and Omicron variants of SARS-CoV-2[24,25], validating the epidemiological relevance of the selected parameters. The corresponding variation in the herd immunity threshold (between 76.2% and 81.4%) underscores the importance of continuous monitoring of clinical variables to project the vaccination coverage targets required in active epidemic scenarios.

It must be emphasized that the biomedical data used correspond to simulated variables generated by the microcontroller for academic and system validation purposes, not to clinical information from real patients[16,17]. This explicitly declared decision allows evaluation of the technical feasibility of the proposed integration in a controlled environment, establishing conceptual and methodological foundations for future developments in digital epidemiological surveillance — a strategic field for strengthening predictive capabilities in public health[18–20].

Table 7 presents a comparative analysis of the main SEIR model variants reported in the literature, evaluating their capacity for real-time updating of the β(t) parameter and their compatibility with the cyber-physical system (CPS) architecture. Classical models with constant β[1,2] or with predefined forcing do not allow incorporation of IoT sensor data. In contrast, the model with simple parametric coupling and its dual extension (temperature + SpO_2_) enable continuous updating of β(t) each time MATLAB retrieves new data from ThingSpeak, thereby closing the CPS feedback loop. More advanced formulations such as the Kalman filter [13] or neural networks[28] are also compatible, but entail greater computational complexity that is not warranted in this conceptual validation phase.

**Table 7.**
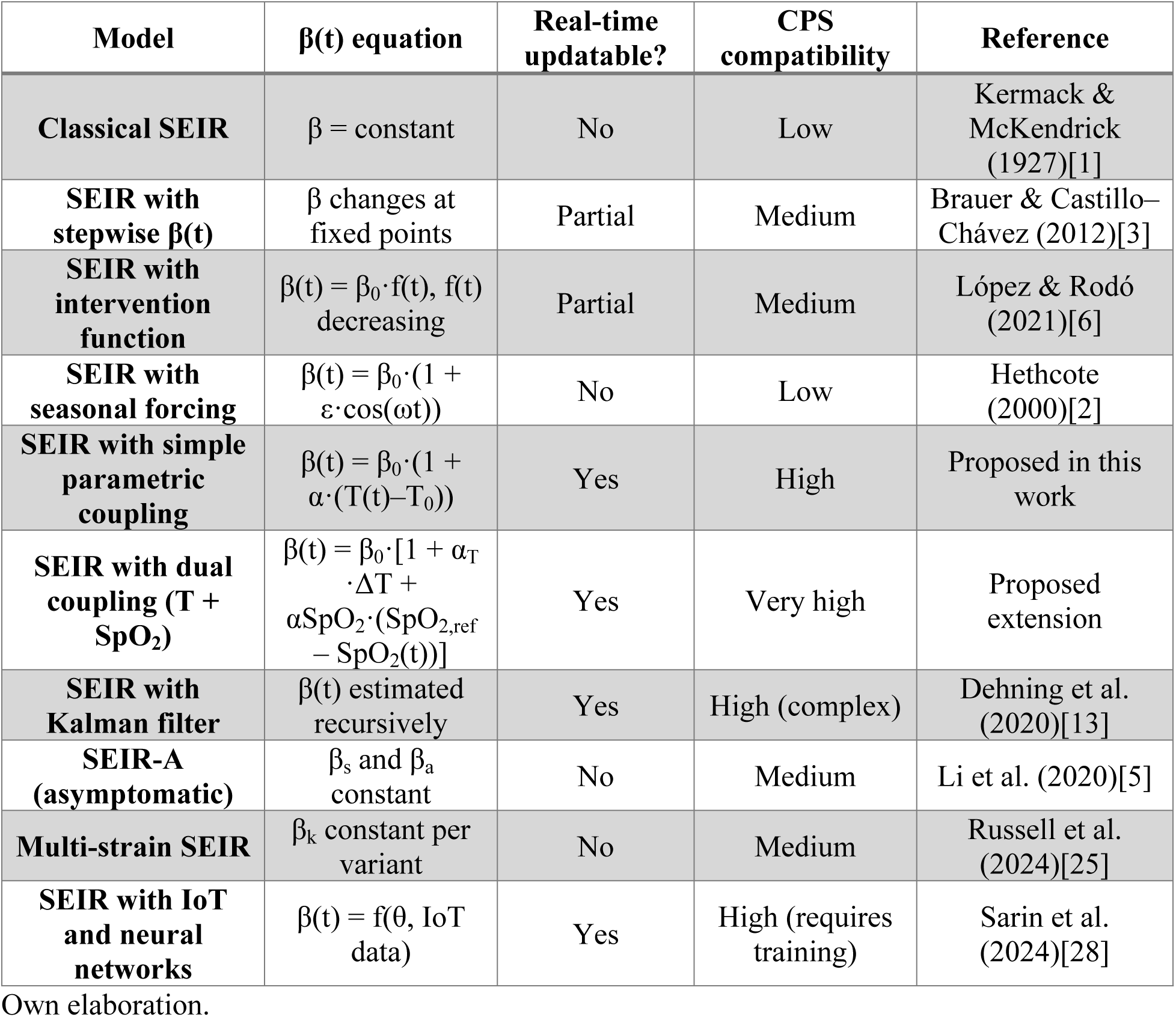
Comparative table: SEIR models and their compatibility with CPS.

| Model | $\beta(t)$ equation | Real-time updatable? | CPS compatibility | Reference |
| --- | --- | --- | --- | --- |
| **Classical SEIR** | $\beta = \text{constant}$ | No | Low | Kermack & McKendrick (1927)[1] |
| **SEIR with stepwise $\beta(t)$** | $\beta$ changes at fixed points | Partial | Medium | Brauer & Castillo-Chávez (2012)[3] |
| **SEIR with intervention function** | $\beta(t) = \beta_0 \cdot f(t)$, $f(t)$ decreasing | Partial | Medium | López & Rodó (2021)[6] |
| **SEIR with seasonal forcing** | $\beta(t) = \beta_0 \cdot (1 + \varepsilon \cdot \cos(\omega t))$ | No | Low | Hethcote (2000)[2] |
| **SEIR with simple parametric coupling** | $\beta(t) = \beta_0 \cdot (1 + \alpha \cdot (T(t) - T_0))$ | Yes | High | Proposed in this work |
| **SEIR with dual coupling (T + $\text{SpO}_2$)** | $\beta(t) = \beta_0 \cdot [1 + \alpha_T \cdot \Delta T + \alpha_{\text{SpO}_2} \cdot (\text{SpO}_{2,\text{ref}} - \text{SpO}_2(t))]$ | Yes | Very high | Proposed extension |
| **SEIR with Kalman filter** | $\beta(t)$ estimated recursively | Yes | High (complex) | Dehning et al. (2020)[13] |
| **SEIR-A (asymptomatic)** | $\beta_s$ and $\beta_a$ constant | No | Medium | Li et al. (2020)[5] |
| **Multi-strain SEIR** | $\beta_k$ constant per variant | No | Medium | Russell et al. (2024)[25] |
| **SEIR with IoT and neural networks** | $\beta(t) = f(\theta, \text{IoT data})$ | Yes | High (requires training) | Sarin et al. (2024)[28] |

Own elaboration.

It is important to note that the proposed coupling function uses body temperature and oxygen saturation as proxy variables of the population epidemic status, not as direct determinants of transmission. From a clinical perspective, fever and hypoxemia are markers of individual disease severity, not of population contagiousness. The relationship between these biomarkers and the transmission parameter β(t) is therefore a design rule of the prototype that allows evaluation of the SEIR model response to quantifiable perturbations, without claiming to establish epidemiological causality between clinical severity and transmissibility.

### Limitations

The present study has several limitations that must be considered when interpreting its results. *Simulated nature of the system and absence of hardware validation.* All biomedical data and the IoT transmission flow were generated computationally; no physical ESP32 hardware was deployed and no real patient data were collected. Consequently, the system performance metrics (latency, transmission success rate, Wi-Fi connection stability, energy consumption) constitute design specifications, not experimental measurements. Operational validation would require physical deployment of the microcontroller with real sensors (MAX30102, MLX90614, or equivalents) and experimental characterization of the acquisition and transmission chain.

*Calibration limited to the baseline SEIR, without calibration of the dual coupling*. The retrospective validation with real Colombian data (Phase 4B) calibrated only the β parameter of the baseline SEIR model (β = 0.1321 days⁻¹, R_0_ = 1.85), without empirically estimating the coefficients of the dual coupling function (αT, αSpO_2_). The scenarios constructed on this calibrated baseline (Fig 8, Panels B and C) illustrate the response of the extended model when β is rescaled to the effective Colombian transmissibility, but do not constitute a validation of the coupling function itself against clinical data. This limitation is methodologically relevant and must be kept in mind when interpreting the projected R_0_ ranges.

*Coupling coefficients chosen by design.* The nominal values αT = 0.05 °C^−1^ and αSpO_2_ = 0.02 %^−1^ were selected as design rules consistent with expected clinical magnitudes, not derived from a statistical inference process on empirical data. The sensitivity analysis presented (Section 3, Phase 5) quantifies the dependence of results on these values and confirms monotonic and predictable behavior, but does not substitute a formal calibration against real biomedical observations.

*Parametric identifiability limitations.* In the current dual coupling formulation, the coefficients αT and αSpO_2_ enter linearly into the same β(t) equation and operate on variables that may be correlated in real patients (fever and hypoxia tend to coexist in severe clinical presentations). Consequently, calibration with limited empirical data could yield multiple combinations of (α_T_, αSpO_2_) fitting the observed dynamics equally well — a parametric identifiability problem. A formal identifiability characterization — through structural and practical identifiability analysis using tools such as DAISY or GenSSI — is a necessary line of work for future versions of the model intended for quantitative calibration.

*Absence of formal uncertainty quantification.* Results are presented as deterministic trajectories obtained with point parameter estimates, without confidence intervals, predictive bands, or formal propagation of uncertainty from parameters to outputs (R_0_, peak infectious proportion, day of peak). Although the univariate and bivariate sensitivity analysis offers a first approximation to the impact of parameter variation, rigorous quantification would require Bayesian approaches (e.g., Markov chain Monte Carlo MCMC over the joint parameter posterior) or uncertainty propagation methods (e.g., polynomial chaos expansions or structured Monte Carlo sampling) that exceed the scope of this proof-of-concept phase.

*Unvalidated causal relationship between individual biomarkers and population transmissibility.* The coupling function uses body temperature and oxygen saturation — individually clinical variables — to modulate a population-level parameter (β(t)). This operation lacks explicit causal support in the epidemiological literature: fever and hypoxemia are markers of individual disease severity, not direct determinants of population contagiousness. The relationship is sustained as a functional design rule of the prototype, not as a validated epidemiological hypothesis.

*Simplifications of the baseline SEIR model.* The model assumes homogeneous mixing and does not incorporate age structure, asymptomatic compartments, or viral variant dynamics (extensions discussed in Table 7). These simplifications are acceptable in an architectural validation phase, but limit the applicability of the system to real epidemic scenarios without the corresponding extensions.

*Size and nature of the simulated population.* The validation was performed with a simulated population of N = 1,000 individuals with fixed parameters derived from the literature. Generalization to real populations (N on the order of 10^6^–10^8^ inhabitants) would require revalidation of the solver’s numerical stability, calibration with observed epidemiological data, and prospective validation against documented outbreaks. The retrospective approach performed in this work (Phase 4B) constitutes a first step toward this validation, but does not substitute it.

## Conclusions

The present work demonstrated the feasibility of designing and implementing a closed-loop cyber-physical architecture integrating a SEIR epidemiological model with simulated biomedical signals through an IoT layer based on the ESP32 and ThingSpeak, implemented entirely in computational simulation.

The baseline SEIR model reproduced epidemic dynamics consistent with the literature, with R_0_ = 4.20 and a temporal lag of approximately 10 days between the peak of exposed individuals E(t) and the peak of infectious individuals I(t) — an emergent property of the joint epidemic dynamics, not equivalent to the individual incubation period (1/σ = 5.2 days). The dual coupling function β(t) = β_0_ · [1 + α_T_ · (T – T_0_) + αSpO_2_ · (SpO_2,ref_ – SpO_2_)] demonstrated its utility as a functional mechanism for the dynamic updating of epidemiological parameters from biomedical signals, generating distinguishable scenarios with R_0_ values between 4.20 and 5.38 depending on the combination of simulated temperature and oxygen saturation.

The sensitivity analysis revealed that the hypoxic sensitivity coefficient (αSpO_2_) exerts greater influence than the thermal coefficient (α_T_) on R_0_, with monotonic and predictable behavior across the entire evaluated parametric space. Simultaneous ±50% variation in both coefficients produced an R_0_ range between 4.79 and 5.96 for the most severe scenario, without qualitatively altering the study conclusions.

The primary contribution of this work is the operational integration of simulated sensing layers, IoT communication, and epidemiological modeling through parametric coupling, in a reproducible platform. The proposed system constitutes a methodological foundation for future developments along four complementary lines: (i) physical deployment of the IoT architecture with real sensors (MAX30102, MLX90614) and experimental characterization of network performance; (ii) empirical calibration of the coupling coefficients (α_T_, αSpO_2_) against clinical cohorts, preceded by structural and practical identifiability analysis; (iii) formal uncertainty quantification via Bayesian schemes (MCMC), Kalman filters for recursive estimation of β(t), or parametric uncertainty propagation methods; and (iv) extensions of the baseline SEIR model toward asymptomatic compartments, age structure, and viral variant dynamics, while maintaining compatibility with the proposed cyber-physical architecture.

## Data Availability

All data, source code, and analysis notebooks underlying the findings of this study are publicly available without restriction in the Zenodo repository under a Creative Commons Attribution 4.0 International (CC BY 4.0) license: https://doi.org/10.5281/zenodo.20027809

## Acknowledgments

The authors thank the Faculty of Engineering at Universidad Cooperativa de Colombia for providing the computational infrastructure used in this study. We also acknowledge the open-data initiative of Our World in Data, which made the retrospective calibration with Colombian COVID-19 surveillance data possible.

## Supporting information captions

**S1 Appendix**. SEIR-IoT cyber-physical architecture with dual parametric coupling for epidemic scenario simulation using synthetic biomedical signals. *(Full manuscript in Spanish)*

**S1 Table.** Sensitivity analysis results: R_0_, peak infectious proportion, and day of peak for 9 combinations of α_T_ (±50%) and αSpO_2_ (±50%) in the Fever+Severe Hypoxia scenario (T = 38.5 °C, SpO_2_ = 88%).

**S1 Fig.** Univariate sensitivity curves: R_0_, peak infectious proportion, and day of peak as a function of α_T_ (upper row) and αSpO_2_ (lower row) for the Normal and Fever+Severe Hypoxia scenarios.

**S2 Fig.** Tornado plot: relative influence of coupling parameters on R_0_ in the Fever+Severe Hypoxia scenario (±50% variation).

**S3 Fig.** Two-dimensional sensitivity: R_0_ and peak infectious proportion as a function of α_T_ and αSpO_2_ for the Fever+Severe Hypoxia scenario (40×40 grid, 1,600 simulations).

**S4 Fig.** Uncertainty bands: I(t) curves for 5 values of αT (left panel) and 5 values of αSpO_2_ (right panel) in the Fever+Severe Hypoxia scenario.

